# Nationally Representative Social Contact Patterns among U.S. adults, August 2020-April 2021

**DOI:** 10.1101/2021.09.22.21263904

**Authors:** Kristin N. Nelson, Aaron J Siegler, Patrick S Sullivan, Heather Bradley, Eric Hall, Nicole Luisi, Palmer Hipp-Ramsey, Travis Sanchez, Kayoko Shioda, Benjamin A Lopman

## Abstract

The response to the COVID-19 pandemic in the U.S prompted abrupt and dramatic changes to social contact patterns. Monitoring changing social behavior is essential to provide reliable input data for mechanistic models of infectious disease, which have been increasingly used to support public health policy to mitigate the impacts of the pandemic. While some studies have reported on changing contact patterns throughout the pandemic., few have reported on differences in contact patterns among key demographic groups and none have reported nationally representative estimates. We conducted a national probability survey of US households and collected information on social contact patterns during two time periods: August-December 2020 (before widespread vaccine availability) and March-April 2021 (during national vaccine rollout). Overall, contact rates in Spring 2021 were similar to those in Fall 2020, with most contacts reported at work. Persons identifying as non-White, non-Black, non-Asian, and non-Hispanic reported high numbers of contacts relative to other racial and ethnic groups. Contact rates were highest in those reporting occupations in retail, hospitality and food service, and transportation. Those testing positive for SARS-CoV-2 antibodies reported a higher number of daily contacts than those who were seronegative. Our findings provide evidence for differences in social behavior among demographic groups, highlighting the profound disparities that have become the hallmark of the COVID-19 pandemic.

## Introduction

The response to the COVID-19 pandemic in the U.S., including the closing of schools, workplaces, and businesses, prompted abrupt and dramatic changes to social contact patterns. In the first months of 2020, reducing social contact was the only measure available to ‘flatten the curve’ and blunt the severity of the early pandemic. A synchronous, near-universal decline in contact rates occurred across countries in North America, Western Europe, and Asia in the Spring and Summer of 2020, with mean daily contacts dropping from 7-26 contacts pre-pandemic to 2-5 contacts per person in the early ‘lockdown’ period. [1] Through Summer and Fall, as restrictions began to ease, contact patterns slowly rebounded. A key inflection point occurred in November 2020, when the first COVID-19 vaccines became available, signaling the start of a massive national vaccination campaign. Widespread vaccination reduced individuals’ risks of infection and led to declining case rates and hospitalizations, contributing to perceptions of reduced pandemic severity and leading to further relaxation of social distancing policies. [2] Characterizing changing social contact patterns across this time period is critical to better understand behavioral drivers of the trajectory of the pandemic and inform ongoing efforts to estimate the impact of interventions. Systematic data collected in other countries has helped to explain the interplay between contact patterns and transmission dynamics [3, 4], but studies of contact patterns in the U.S. during this period draw primarily from convenience samples which survey unrepresentative segments of the population. [5, 6]

While some studies have reported on changing contact patterns throughout the COVID-19 pandemic in the U.S. [1], few have reported on differences in contact patterns among key demographic groups. This is particularly important since the burden of the COVID-19 pandemic has disproportionately fallen on low-income and minority populations, with a heavier burden of COVID-19 cases and deaths in low-income and minority populations. [7-10] To date, research has suggested that such disparities reflect limited capacity for these groups to markedly change their contact patterns to protect themselves and their communities from SARS-CoV-2 infection due to social and structural factors. [11-13] Indeed, studies on neighborhood-level mobility support the notion that certain demographic groups were more likely to self-isolate than others during the early pandemic. [14, 15] Disparities in the impact of COVID-19 persisted as vaccines became available. Inequities in vaccine access and uptake have resulted in suboptimal vaccination rates in many U.S. communities. [16-18] A person’s vaccination status, as well as the vaccination coverage in their broader community, influences the real and perceived risk of infection, shaping differences in social contact patterns across demographic groups as the pandemic continues.

Data are needed on contact patterns over the period of the COVID-19 pandemic in the U.S.; ideally these data should be both nationally representative and investigate differences by demographic groups. Such data are critical to illustrate a national picture of contact patterns driving the changing epidemiology of COVID-19 and provide robust data on the behavioral patterns that may be perpetuating inequities in the impact of the pandemic. Specifically, unbiased estimates of contact will be useful in parameterizing mechanistic models of infectious disease transmission, informing underlying assumptions about the rate at which individuals come into contact with one another which are necessary to accurately simulate spread of infection and the potential impact of interventions. In particular, it is important to capture contact patterns in the period during which restrictions began to be substantially relaxed but before the availability of vaccination in the general population and after in order to guide models aiming to understand the role of vaccines in altering population-level transmission dynamics. With infectious disease models increasingly informing both domestic and global policy, it is essential that the data underlying models maintains fidelity to rapidly changing contact patterns, which represents one of the key sources of uncertainty in COVID-19 transmission models.

We conducted a national probability survey of US households (The COVIDVu Study) with the primary aim to generate nationally representative estimates of the cumulative incidence of SARS-CoV-2 infection. [19-21] As a part of this study, we collected information on social contact patterns during two time periods: August-December 2020 (before widespread vaccine availability) and March-April 2021 (during national vaccine rollout). We calculate nationally representative contact rates for the U.S. by demographic group and assess the relationship between contact rates and SARS-CoV-2 serostatus in both periods.

## Methods

### Sampling

Sampling methods for the COVIDVu study have been previously described. [19] Briefly, we used a national address-based household sample, which was chosen to be representative on the basis of race/ethnicity, age, gender, education level, household income, region of residence, and home ownership. To achieve a total sample of at least 4,000 responding households, a total of 39,500 addresses were sampled. To ensure adequate participation of key groups, we oversampled households in census tracts with >50% Black residents and households with surnames likely to represent Hispanic ethnicity. [22] One household member aged ≥18 years was randomly selected and offered participation in the study. Baseline surveys were conducted from August-December 2020 and a follow-up survey was conducted in March – April 2021. The baseline period coincided with the relaxation of many pandemic restrictions that had lasted through the Summer of 2020 and the subsequent tightening of restrictions as wintertime case counts began to rise in November and December. The follow-up period coincided with the start of widespread COVID-19 vaccine availability and the fall of case counts after the wintertime surge (Supplemental Figure 1).

To adjust for non-response, we calculated sampling weights. Weights were used to estimate key parameters that represent the non-institutionalized, housed adult population in the U.S. The COVIDVu study was approved by the Emory University Institutional Review Board (STUDY00000695).

### Survey and specimen collection

Consenting participants completed an online survey, which collected the age, sex, race, ethnicity, income, household size, and occupation of participants. We also included questions about person-to-person contacts on the previous day, which we adapted from previous, similar surveys. [6, 23, 24] We defined a physical contact as any contact involving physical touch, such as a handshake, hug, or kiss and a non-physical contact by an interaction in which the participant was within 6 feet of the other person and exchanged three or more words. For each contact reported in the survey, participants were asked to record the age of the contact and the location where contact occurred (home, work, school, or other). Occupation categories were adapted from those used in the 2018 American Community Survey and based on the North American Industry Classification System. [25]

Participants self-collected and returned a dried blood spot (DBS) specimen to a central laboratory via prepaid mailer. DBS specimens were tested using the BioRad Platelia Total Antibody test that targets the nucleocapsid protein (i.e., IgA, IgM, IgG; BioRad, Hercules, California). [26, 27]

### Statistical Analysis

We restricted our analysis to participants who completed the baseline and follow-up surveys and returned DBS specimens from each period with valid total Ig result. Where missing, we imputed responses for all variables involved in calculation of weights, including age, gender, race/ethnicity, and income. [19, 28] Using the survey weights, we calculated the mean and median daily number of contacts reported in each time period, overall and by age, sex, location of contact, race/ethnicity, income, occupation, and SARS-CoV-2 serostatus. We performed paired two-sample t-tests and Kolmogorov-Smirnov tests for the difference in contacts reported at baseline and follow-up, which assess differences in the mean and overall distribution, respectively, of highly skewed data. We assessed the relationship between number of contacts and serostatus adjusting for age and race/ethnicity, which we have previously found to be associated with serostatus. [21] For comparability with analyses of social contact from other countries, we truncated data at 50 contacts per day per contact age group to report total contacts [4], though we explored other cutoffs to assess how the impact on our findings. All analysis was conducted in R version 3.5.3 and all code is available on GitHub at https://github.com/kbratnelson/covidvu-socialcontact. A dataset including contact variables and key sociodemographic variables, as well as survey weights, is also available on GitHub.

## Results

39,500 registration materials and kits were mailed to sampled US households. 4,654 (12.6% of sampled households) returned a baseline survey and a DBS specimen with a valid total Ig result during the period August 9-December 8, 2020. Of these, 3,112 (66.9% of participants in the baseline survey) returned a follow-up survey and a DBS specimen with a valid total Ig result during the period March 2-April 18, 2021.

Overall, the mean number of daily contacts was 13.9 (IQR: 2, 10) at baseline and 14.5 (IQR: 2, 11) at follow-up (p of two-sample t-test = 0.552). (Table 1) Physical contacts increased slightly from 5.9 (IQR: 1, 4) to 7.8 (IQR: 0, 5) contacts (p=0.322), and non-physical contacts stayed similar (10.2 (IQR: 0, 5) at baseline and 9.6 (IQR: 0, 6) at follow-up, p=0.722). Most contacts in both survey periods were reported at work (65% at baseline and 54% at follow-up), with fewer contacts reported at home (17% at baseline and 21% at follow-up) and other locations (17% at baseline and 22% at follow-up); the fewest contacts were reported at school (1% at baseline and 3% at follow-up). (Figure 1) (Of note, nearly all participants were adults, so contacts at school were not expected to be common.) While the distribution of contacts in all locations was skewed, this skew was particularly apparent among workplace contacts (90^th^ percentile of the distribution: 16 and 15 for baseline and follow-up, respectively).

**Table 1.**
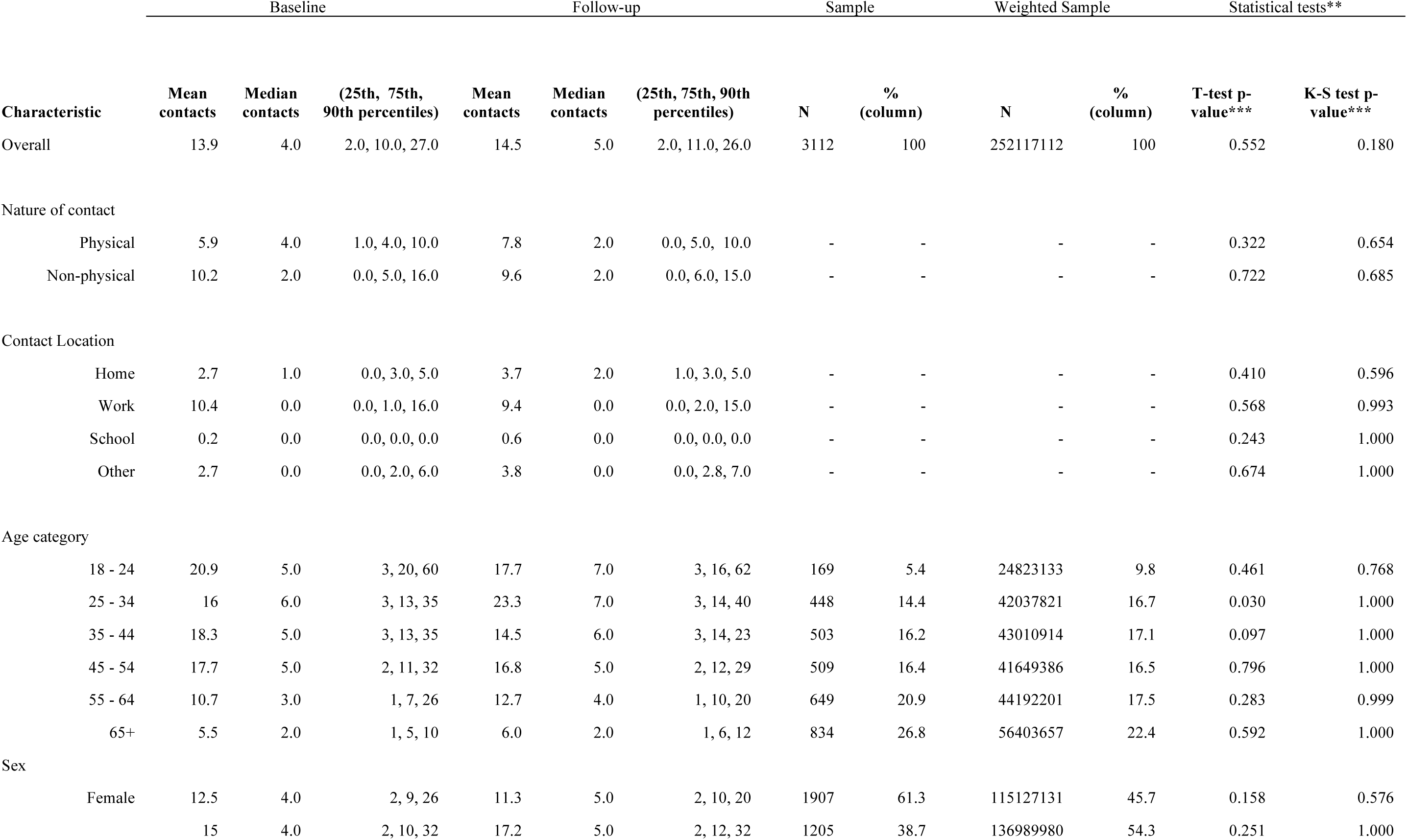
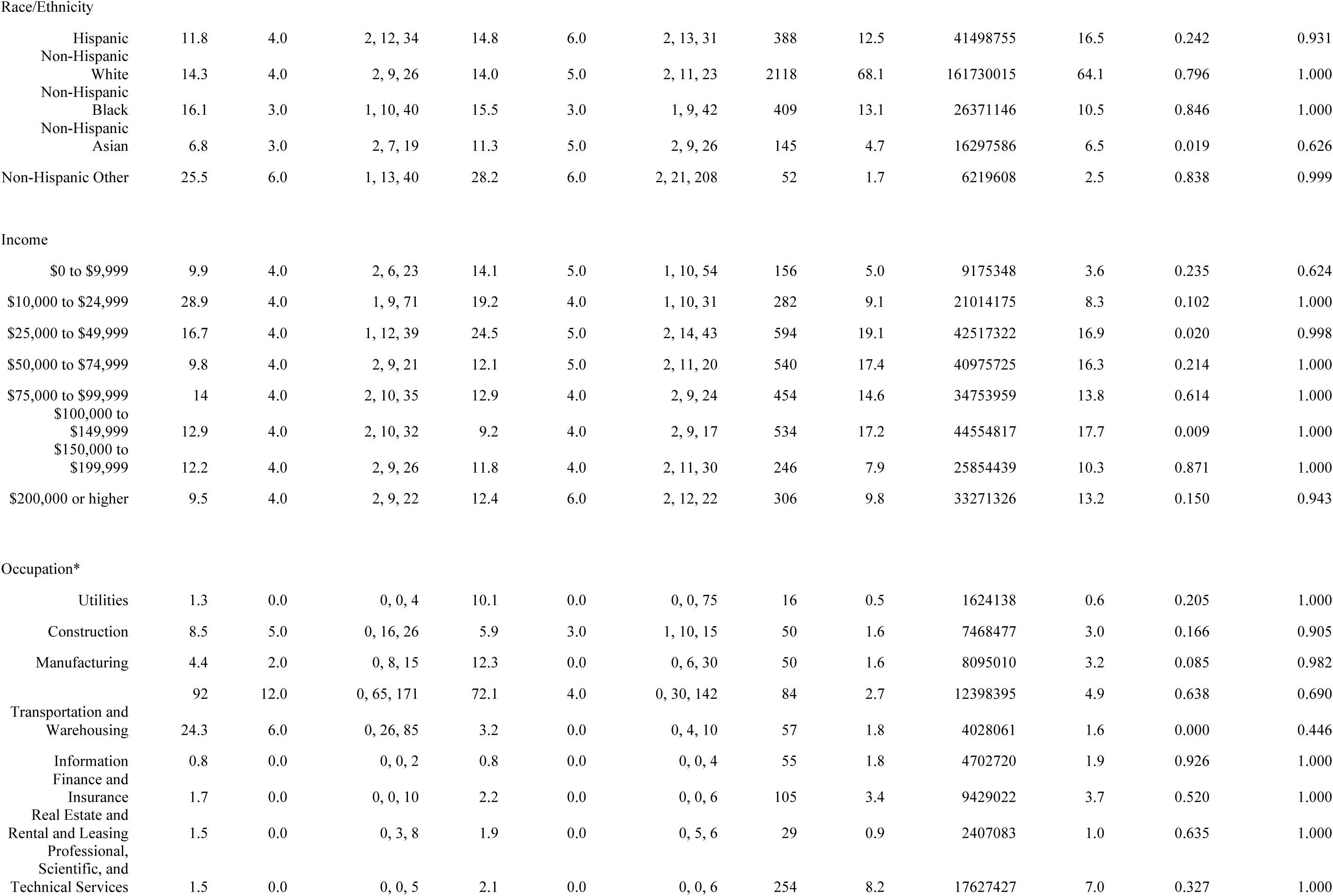
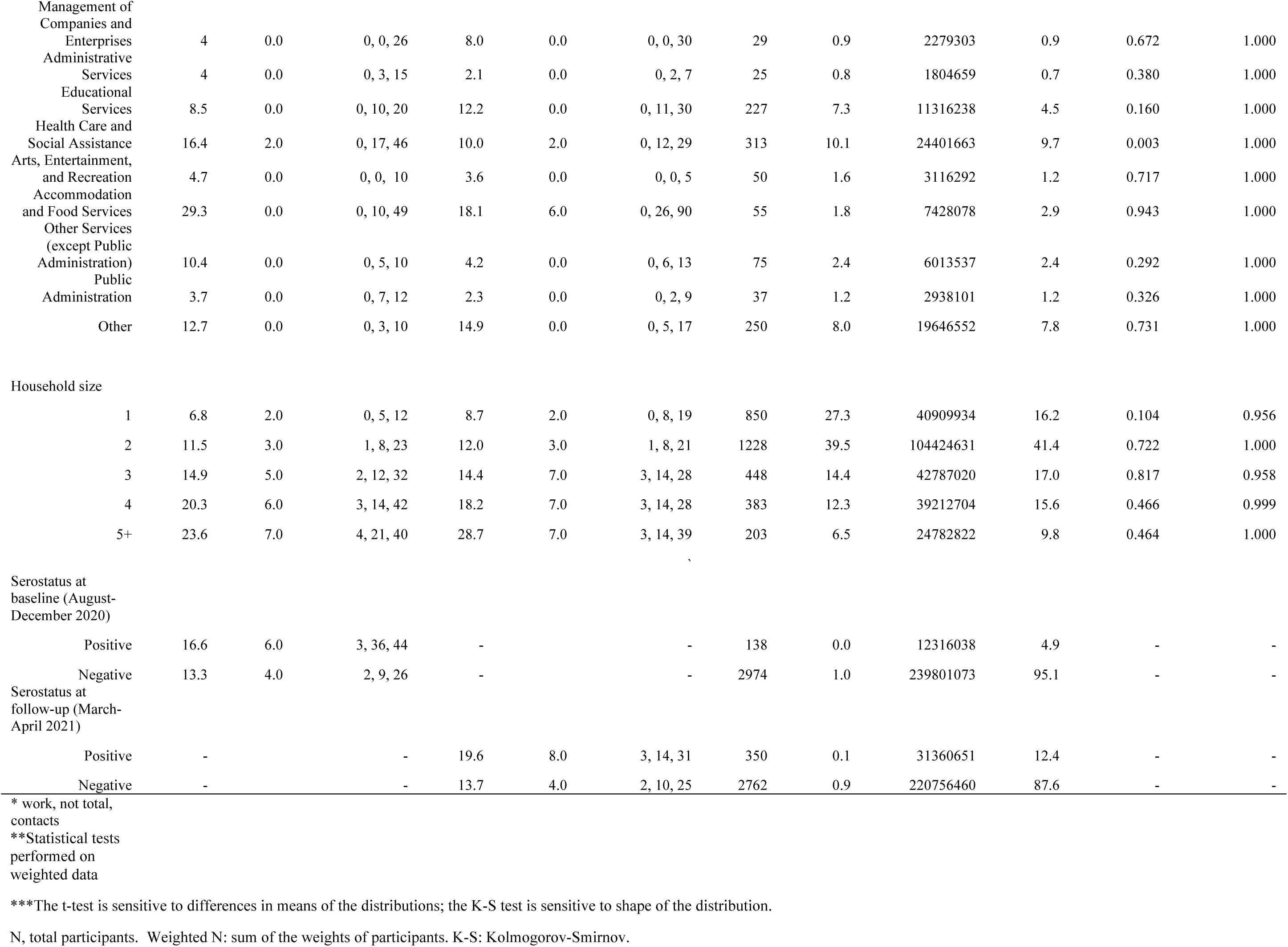
Mean, median, and quantiles of reported contacts per day by contact location, type, demographic group, and serostatus for a probability sample of 3,112 households representing the population aged ≥18 years, United States, 2020-2021

**Figure 1.**
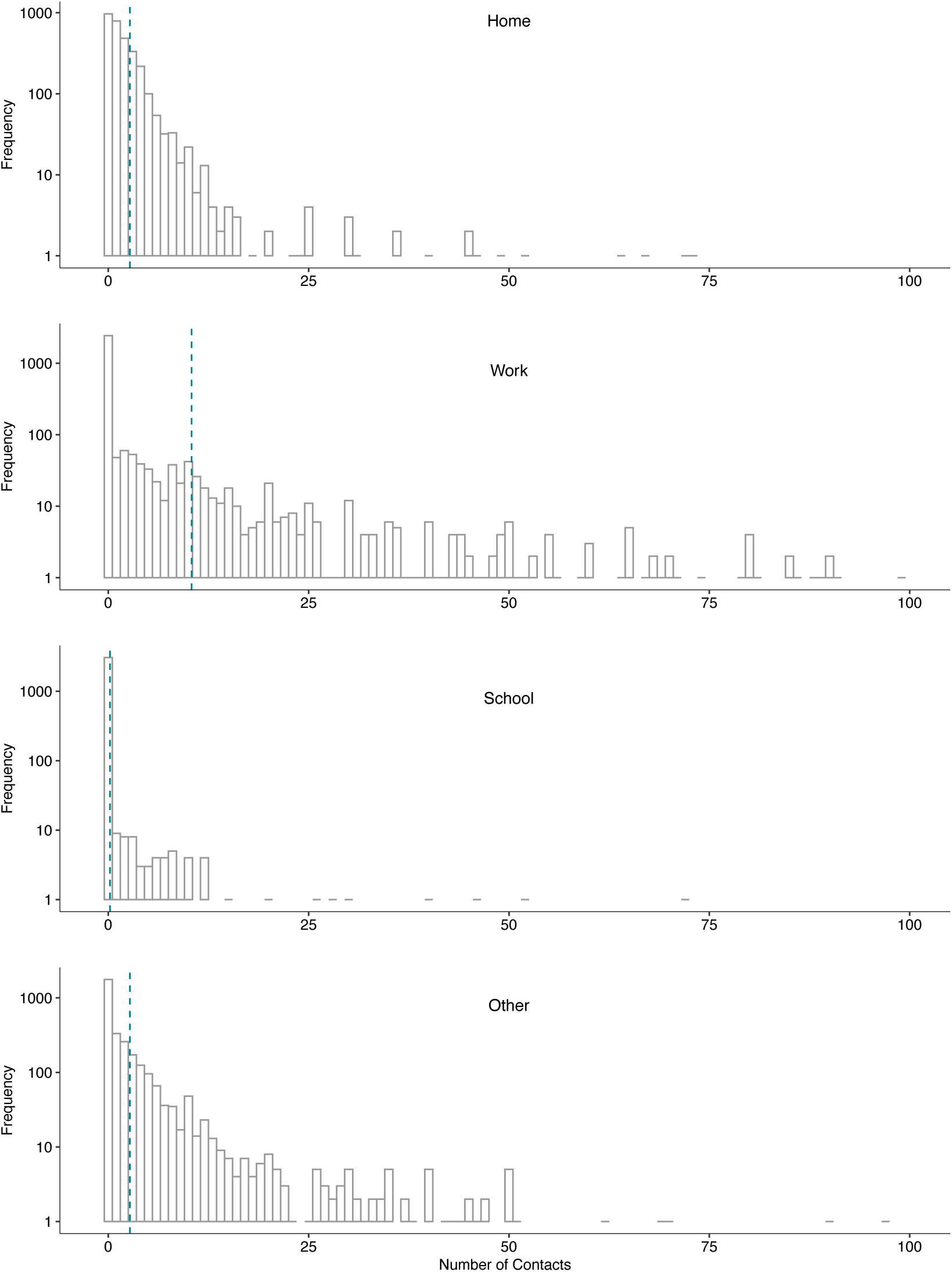
Frequency of reported contacts per day by location of contact for a probability sample of 3,112 households representing the population aged ≥18 years at study baseline, United States, August-December 2020 Contacts reported in each location are truncated at 100 to display the lower end of the distribution for each plot. Vertical dotted line shows the mean for each location (as reported in Table 1).

Trends in contact varied by age group (Figures 2 and 3). The number of contacts did not substantially change among 18-24 year-olds (p=0.461), 35-44 year-olds (p=0.097), and 45-54 year-olds (p=0.796) from baseline to follow-up; comparatively, contact increased among 25-34 year-olds (mean of 16 contacts at baseline and 23.3 contacts at follow-up, p=0.030). Contact rates among the elderly (65+) were the lowest of all age groups and remained similar from baseline to follow-up (mean of 5.5 and 6.0 contacts at baseline and follow-up, respectively, p=0.592). The number of contacts among men were generally higher than among women and were similar from baseline to follow-up (15.0 to 17.2, p=0.251), contacts reported among women were also similar between the two surveys (mean of 12.5 and 11.3 at baseline and follow-up, respectively, p=0.158).

**Figure 2.**
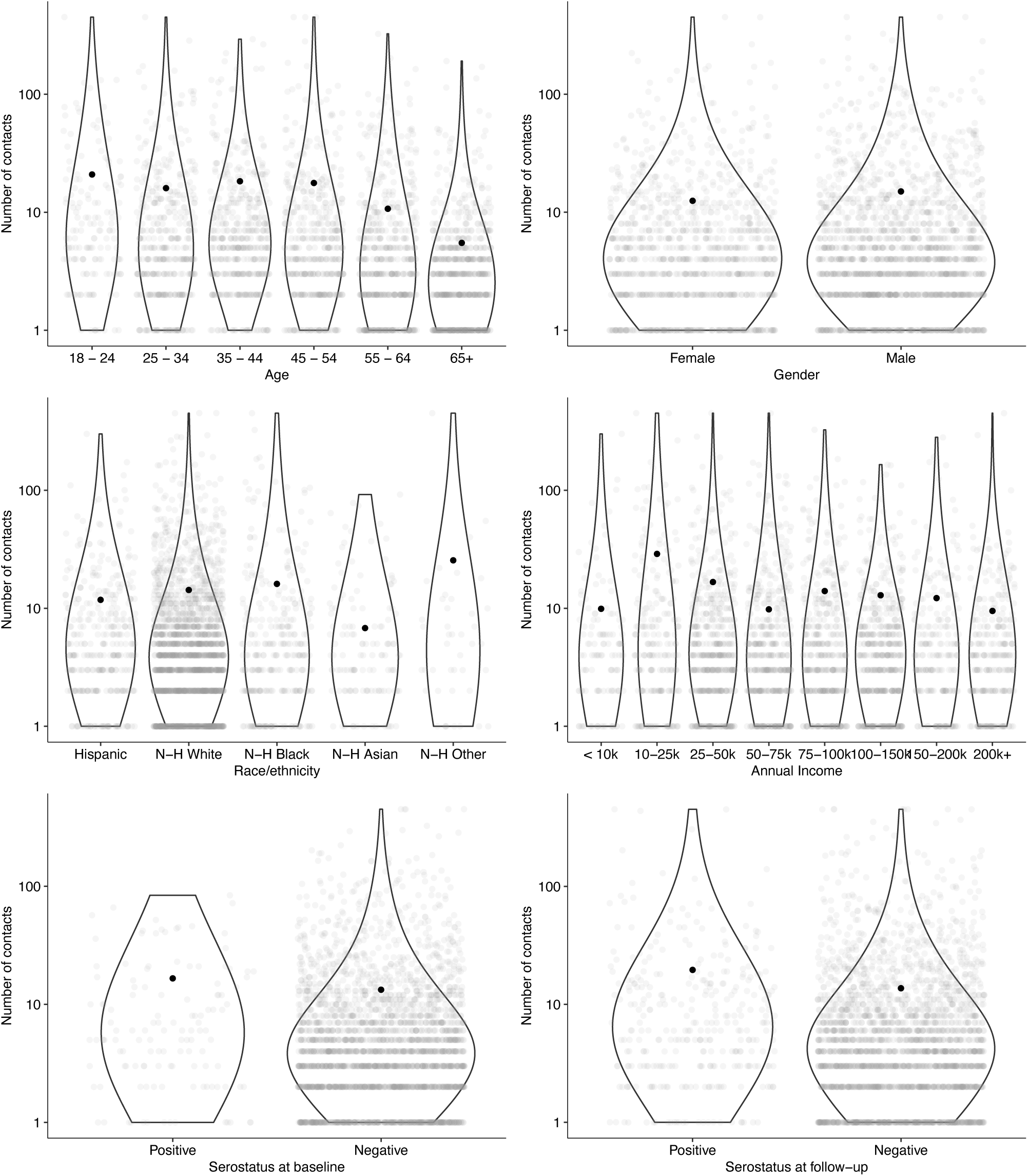
Mean reported contacts per day by demographic group and serostatus for a probability sample of 3,112 households representing the population aged ≥18 years at study baseline, United States, August-December 2020 Violin plots show the distribution of the non-zero numbers of contacts reported by group and grey dots indicate each individual data point. Black dots show the mean for each group (as reported in Table 1).

**Figure 3.**
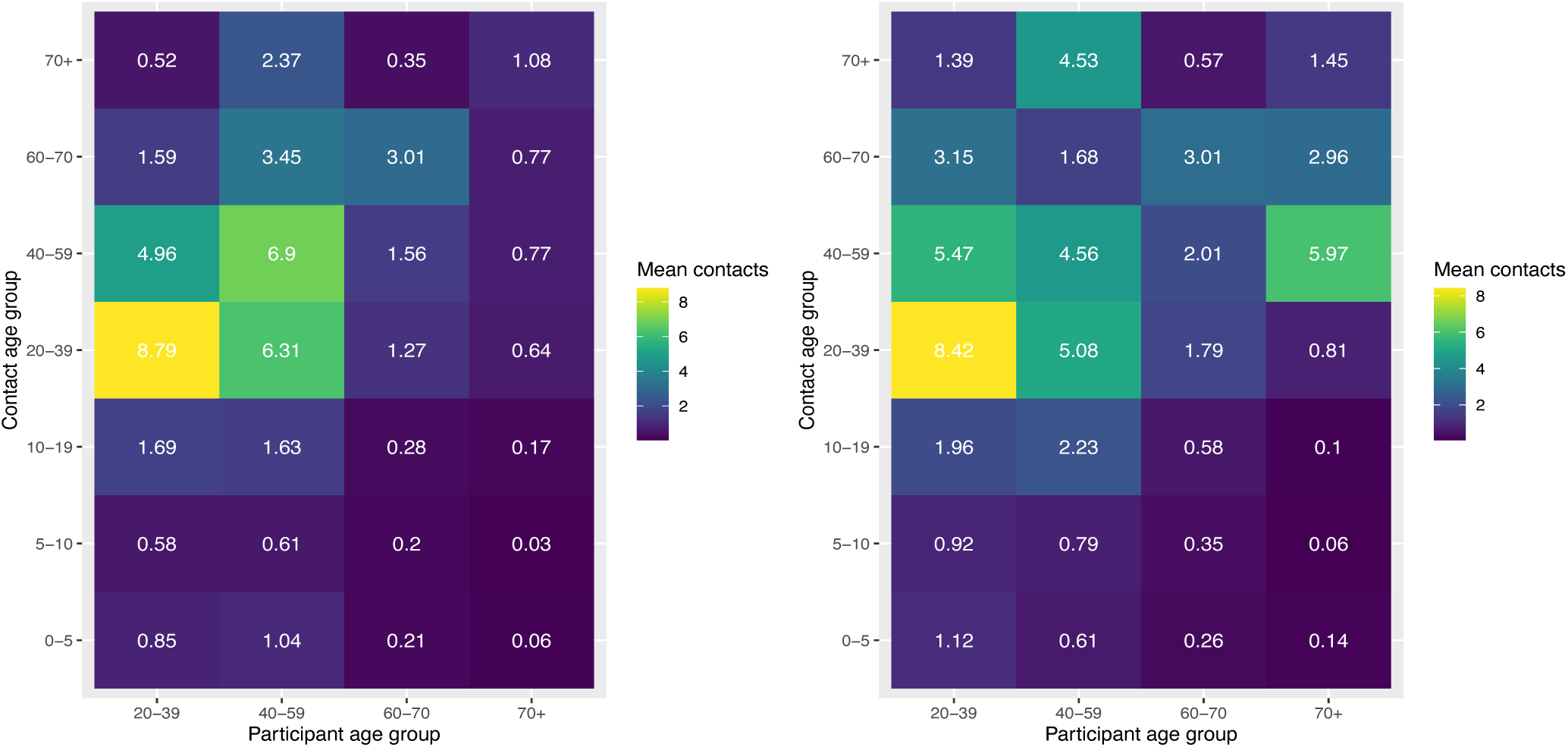
Mean contact rates per day by age group for a probability sample of 3,112 households representing the population aged ≥18 years at study baseline, United States, August 2020-April 2021 Mean number of total contacts (physical and non-physical) reported among participants by age group at baseline (left) and at follow-up (right), accounting for survey weights. Children were not included in our study but participants could report contact with children, so contact matrices are asymmetric.

Contact patterns were distinct across socioeconomic and racial groups in both surveys (Figure 2), but showed little change from baseline to follow-up. Those identifying as Asian reported the lowest contact rates (6.8 and 11.3 mean contacts at baseline and follow-up, respectively, p=0.019), and those who identified as non-White, non-Black, non-Asian, and non-Hispanic (‘Other’) reported the highest contact rates (baseline mean: 25.5, follow-up mean 28.2, p=0.838). Number of reported contacts was highest among those reporting annual incomes from $10,000-$49,999. The number of contacts reported at work varied substantially by industry, with jobs in retail (mean 92 contacts), accommodation and food service (mean 29 contacts at baseline), transportation (mean 24 contacts at baseline), healthcare (mean 16 contacts at baseline), manufacturing (mean 12.3 contacts at follow-up), and education (mean 12 contacts at follow-up) accounting for the highest levels of contact.

Participants with a positive serostatus had slightly higher number of contacts than those with a negative serostatus in both periods (16.6 compared to 13.3 contacts at baseline and 19.6 compared to 13.7 contacts at follow-up). (Figure 2) Those who were seronegative at both surveys reported a lower mean number of contacts than those who were seropositive at either survey (13.4 and 16.8 contacts at baseline and follow-up, respectively). Those who seroconverted from baseline to follow-up reported a higher number of contacts at both surveys (17.0 and 18.2 contacts) than those who did not (13.6 and 14.1 contacts). Number of contacts reported was not associated with serostatus at either survey after adjusting for age group and race/ethnicity.

We chose to truncate contacts at 50 per day per contact age group (9 age groups x 50 contacts = total of 450 contacts) to enable comparison with other pandemic-focused studies of contact. Moreover, we reasoned that the nature of some occupations could reasonably lead to several hundred contacts a day. Very few participants reported above the 50 per day per group limit of contacts (5, or 0.2% of participants, at baseline and 10, or 0.3% of participants, at follow-up). (Supplemental Table 1) When we used a truncation limit of 29 total contacts per day, consistent with other previous studies of contact [23], we found qualitatively similar results to those reported (mean 7.7 contacts (IQR: 2, 10) at baseline to 8.1 contacts (IQR: 2, 11) at follow-up. A moderate number of participants reported more than 29 contacts (220 participants (7%) at baseline and 249 participants (8%) at follow-up). In general, those reporting a very high number of contacts reported them at work or at ‘other’ locations.

## Discussion

Overall, national contact rates in Spring 2021 were similar to those in Fall 2020, with most contacts in both surveys reported at work. The number of contacts reported was not uniform across groups, with those identifying as non-White, non-Black, non-Asian, (‘Other’ race) and non-Hispanic reporting high rates of contact relative to other racial and ethnic groups. Contact rates were highest among those with lower incomes and in specific occupational categories, including retail, hospitality and food service, and transportation. While the number of contacts reported were mostly similar from baseline to follow-up, younger adults (aged 25-34) reported higher numbers of contacts at follow-up as compared to baseline. Finally, we found that those testing positive for SARS-CoV-2 antibodies reported a higher number of daily contacts than those who were seronegative. Collectively, these findings provide robust empirical evidence for differences in social behavior among demographic groups, highlighting the profound disparities that have become the hallmark of the COVID-19 pandemic.

The ‘opening up’ of much of the U.S. in mid-2020 saw the lifting of restrictive social distancing measures put into place across much of the country at the beginning of the epidemic. Social contact reached a nadir in Spring 2020, began to rebound by Fall 2020, and according to our findings stayed largely consistent through Spring 2021. The consistency of overall contact rates across these two periods is ostensibly the result of a complex interplay of factors. The expiration of many local mitigation policies in late Summer 2020 [29] likely prompted increases in contact, before the severe wintertime ‘wave’ arrived in November caused contact rates to decline once again. Optimism about the introduction of vaccines in early 2021 gradually began to counterbalance the caution prompted by rising case counts in winter [2]. Together, these factors likely led to largely consistent social contact rates across the two survey periods.

Although pre-pandemic data on social contact is limited, available data suggest that contact rates may have rebounded to close to pre-pandemic levels, estimated at 12-14 contacts per day for the average adult. [30, 31] Comparisons with pre-pandemic contact data are not straightforward, given that previous studies have use different methods to collect contact data and no previous, nationally representative surveys such as this one have been performed. [32] However, we suspect that while overall numbers of contacts may have rebounded, the nature of contact (i.e., physical vs. non-physical, masked vs. unmasked) has changed and, moreover, occurred unevenly among demographic groups. Of note, this return to pre-pandemic contact levels is in contrast to data from European countries, which has shown that as of early 2021 (UK) and mid-2020 (Belgium), contact rates had yet to return to pre-pandemic levels. [33, 34]

Our finding that social contact rates varied by demographic group in the U.S. has been shown in smaller, less broadly representative studies [5] We found high contact rates among the youngest age group, 18-24 year-olds, consistent with other data showing that this group typically reported lowest prevalence of mitigation behaviors. [35-37] We also found that adults 24-35 increased their contact rates the most from baseline to follow-up, which may reflect the opening of schools and workplaces over this period, and later, increased contacts after becoming partially or fully vaccinated. Minority populations and those with lower incomes reported high mean rates of contact. Together with the fact that the majority of contacts at both baseline and follow-up were reported in the workplace, this is consistent with reports that lower-income and minority groups were more likely to be classified as ‘essential’ workers, a designation which may have required them to continue working in-person or to return to work shortly after the most strict social distancing restrictions were lifted and prior to our baseline survey. [13] At least some of this disparity in contact rates is likely due to inflexible and unstable employment situations that are less likely to allow for remote work. Differential contact rates by job type were expected but striking in magnitude. We found contact rates up to ten times the population average in occupations which involve working environments that place employees in close contact with many other people, such as retail, transportation, and food processing. Notably, workplaces in general are an important source of infection and transmission risk for respiratory infections [38-40] and the industries associated with high contact rates in our survey were the settings of several highly publicized outbreaks in 2020. [41, 42] Ensuring that effective prevention measures are enacted in workplaces, where exposure rates are high, may disproportionately reduce infection risk.

Finally, we found that seropositive individuals were more likely to report higher contact rates at both survey rounds. Many studies have reported on risk factors for infection related to social environment, which have included poverty and crowding. [43-45] We coupled behavioral information with serological status in a nationally representative sample to show that higher rates of contact are indeed linked with previous infection status. However, we did not find that contact rate was associated with serostatus after adjusting for age and race/ethnicity, suggesting a more central role for demographic factors over purely behavioral metrics such as contact rate. Linking information on mitigation behaviors and serostatus will be important in understanding epidemic dynamics as population-level immunity increases, which are predicted to be characterized by highly localized outbreaks in space and time driven by ‘pockets’ of susceptible persons. [46, 47]

The availability of nationally representative data on contact patterns by demographic group fills an important gap in efforts to accurately forecast and explain the trajectory of the COVID-19 pandemic. Traditionally, infectious disease models have rarely incorporated social factors, though they are instrumental in determining disease risk. This is partially because of limited availability of data to properly formulate models that track many population groups, but also because average population models are often favored for their simplicity and greater ease of computation. However, accounting for exposure differences between groups is a critical component of accurately modeling disease dynamics by population segment. To date, most models that have aimed to explain differences in the burden of cases and deaths by demographic group have used proxies of contact, such as mobility measured by mobile phone location data, to represent the frequency of person-person interactions rather than data on direct interactions. However, recent work has shown that during ‘lockdowns’, mobility was likely to be a poor indicator of contact patterns, calling into question the validity of this assumption. [48] Data on contact patterns by race, ethnicity, socioeconomic group, and occupation provides the necessary granularity to investigate disease dynamics driving sociodemographic inequities in disease burden. Indeed, modeling studies that have investigated these questions have noted the lack of longitudinal social contact surveys that would provide data to allow proper parameterization of models. [49] Systematically collected data that is finely stratified by demographic group has to date been a major barrier to COVID-19 research efforts. [50] Collecting and making this data available to the modeling community can support the development of more accurate models to inform public health policymaking on continued use of vaccines as well as non-pharmaceutical interventions. As vaccine rollout continues and transmission becomes increasingly concentrated among unvaccinated and undervaccinated populations, understanding between and within-group interactions will be particularly critical to track trends in transmission over time and to understand the potential utility of targeted interventions to mitigate population-level risk.

There were several limitations to this study. First, our response rates were low (<15%) but standard for address-based surveys. For certain groups at risk of being underrepresented by our initial survey responses, we intentionally oversampled to ensure that such groups were adequately represented, and survey weights were used to help ensure the sample represented the underlying population. Second, contact patterns may have changed over the period spanned by our survey (four months in the baseline survey, one month in the follow-up). In addition, our surveys were conducted before and after the winter holidays, which likely saw transient increases in travel and contact rates; we could not capture these changes with our surveys. Third, differences in contact patterns on the state and local levels are likely to be significant due to variation in the nature of and adherence to distancing and other mitigation policies as well as differences in the speed of vaccine rollout and eligibility criteria. The national focus of our survey limits the inferences we can make about contact patterns at a finer geographic scale. With respect to the contact survey, we asked participants to record contacts from one day (the day prior to completing the survey) and we do not know to what extent this day might represent general patterns of contact. Moreover, this prevents an analysis of whether reported contacts were unique on that day, or repeated across days. Generally, a higher number of unique contacts would represent higher infection and transmission risk. Fourth, we do not have information on mitigation measures relevant for each contact, which impact the likelihood of transmitting infection during an interaction. Lastly, the age range of the surveyed population limits the generalizability of these results to only adults, as there were not a sufficient number of participants under 18 to make conclusions about contact rates among younger people. This also limits our view into school-based contact, which is potentially an important setting of transmission.

As the pandemic continues, continued collection of data on behavioural indicators by key demographic groups can support the development of detailed COVID-19 models that accurately represent disease dynamics in varied sociodemographic contexts. Continued, systematic collection and analysis of social contact data is critical to understand the past and future trajectory of the epidemic as well as explain differences in disease burden by population segment and through time. Longitudinal social contact data is particularly powerful alongside seropositivity estimates that are finely stratified by demographic group, and can further support efforts to properly fit dynamic models aiming assess interventions to mitigate the continuing impacts of the pandemic.

## Data Availability

Data are available upon request from the authors. All code is available on GitHub at https://github.com/kbratnelson/covidvu-socialcontact.

## Supplemental Material

**Supplemental Table 1.**
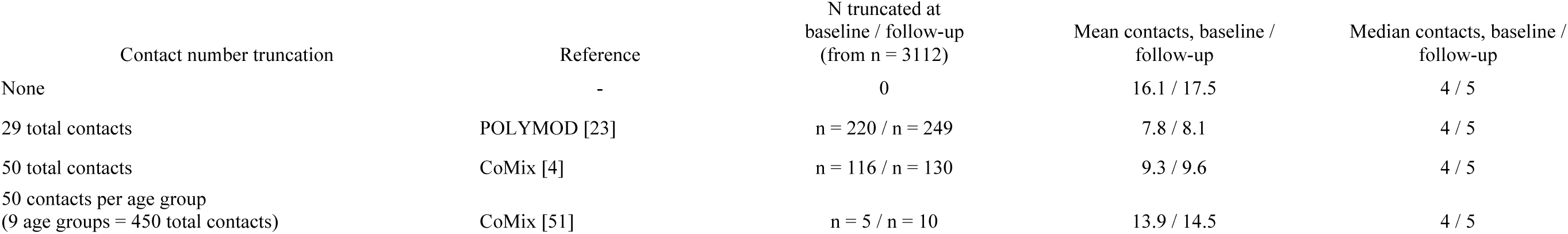
Considering different cutoffs for reported contacts

**Supplemental Figure 1.**
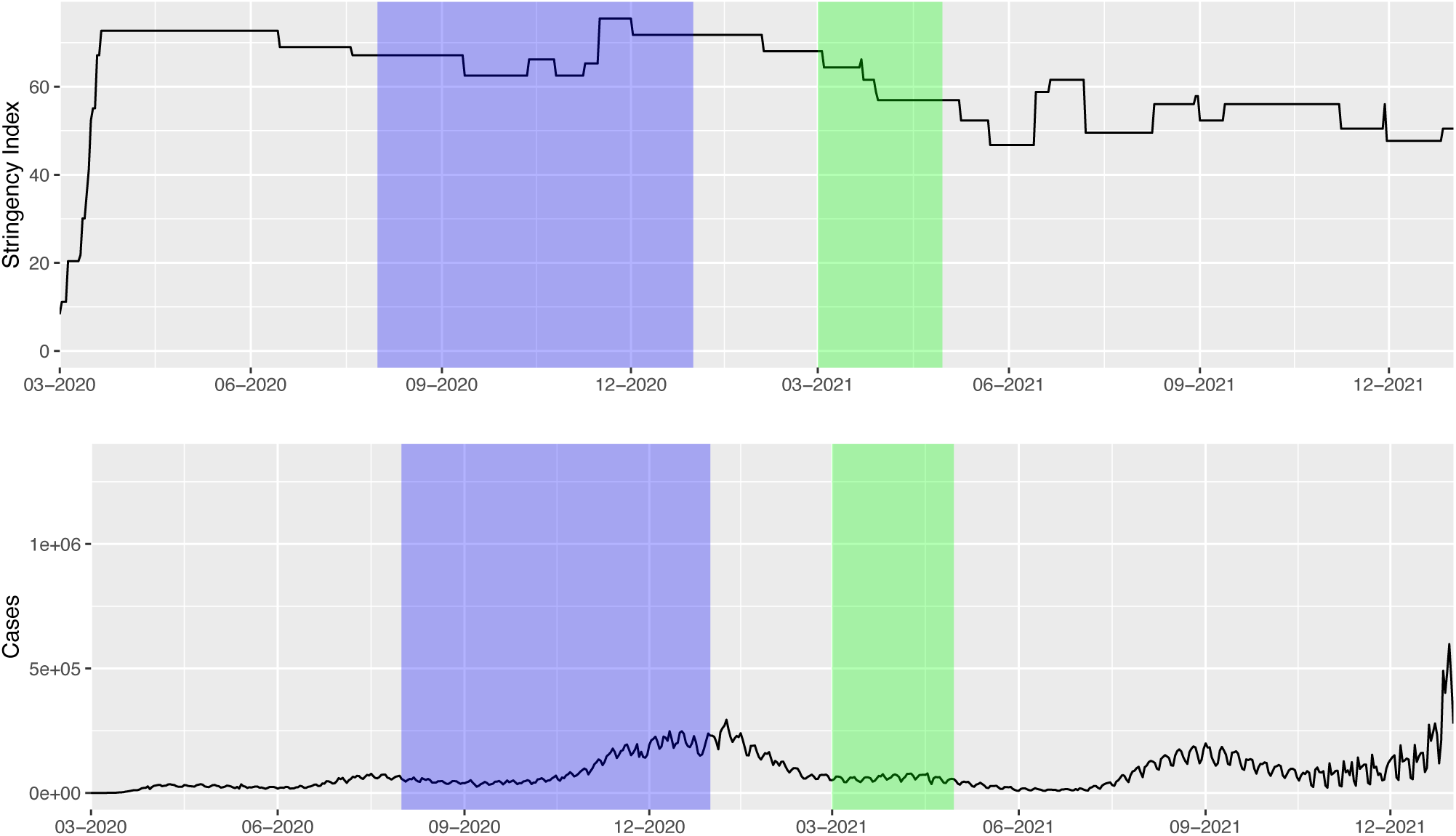
Oxford COVID-19 Government Response Tracker (OxCGRT) Stringency Index and Reported COVID-19 Cases in the U.S. The Oxford COVID-19 Government Response Tracker (OxCGRT) provides a systematic set of cross-national, longitudinal measures of government responses from 1 January 2020. The project tracks national governments’ policies and interventions across a standardized series of indicators and creates a suite of composite indices to measure the extent of these responses.[52] The blue band shows the timing of the baseline survey and the green band shows the timing of the follow-up survey. U.S. COVID-19 cases are plotted using data from the Centers for Disease Control and Prevention COVID data tracker.

